# Incidence, clinical outcomes, and transmission dynamics of hospitalized 2019 coronavirus disease among 9,596,321 individuals residing in California and Washington, United States: a prospective cohort study

**DOI:** 10.1101/2020.04.12.20062943

**Authors:** Joseph A. Lewnard, Vincent X. Liu, Michael L. Jackson, Mark A. Schmidt, Britta L. Jewell, Jean P. Flores, Chris Jentz, Graham R. Northrup, Ayesha Mahmud, Arthur L. Reingold, Maya Petersen, Nicholas P. Jewell, Scott Young, Jim Bellows

**Affiliations:** Division of Epidemiology & Biostatistics, School of Public Health, University of California, Berkeley, Berkeley, California 94720; Division of Infectious Diseases & Vaccinology, School of Public Health, University of California, Berkeley, Berkeley, California 94720; Center for Computational Biology, College of Engineering, University of California, Berkeley, Berkeley, California 94720; Division of Research, Kaiser Permanente, Oakland, California 94612; Health Research Institute, Kaiser Permanente Washington, Seattle, Washington 98101; Center for Health Research, Kaiser Permanente Northwest, Portland, Oregon 97227; MRC Centre for Global Infectious Disease Analysis, Abdul Latif Jameel Institute for Disease and Emergency Analytics, Imperial College London, London, United Kingdom; Department of Infectious Disease Epidemiology, Imperial College London, London, United Kingdom; The Care Management Institute, Kaiser Permanente, Oakland, California 94612; Department of Demography, University of California, Berkeley, Berkeley, California 94720; Department of Medical Statistics, London School of Hygiene & Tropical Medicine, London, United Kingdom; The Permanente Federation, Kaiser Permanente, Oakland, California 94612

## Abstract

**Background:** The United States is now the country reporting the highest number of 2019 coronavirus disease (COVID-19) cases and deaths. However, little is known about the epidemiology and burden of severe COVID-19 to inform planning within healthcare systems and modeling of intervention impact.

**Methods:** We assessed incidence, duration of hospitalization, and clinical outcomes of acute COVID-19 inpatient admissions in a prospectively-followed cohort of 9,596,321 individuals enrolled in comprehensive, integrated healthcare delivery plans from Kaiser Permanente in California and Washington state. We also estimated the effective reproductive number (*R*_*E*_) describing transmission in the study populations.

**Results:** Data covered 1277 hospitalized patients with laboratory- or clinically-confirmed COVID-19 diagnosis by April 9, 2020. Cumulative incidence of first COVID-19 acute inpatient admission was 10.6-12.4 per 100,000 cohort members across the study regions. Mean censoring-adjusted duration of hospitalization was 10.7 days (2.5-97.5%iles: 0.8-30.1) among survivors and 13.7 days (2.5-97.5%iles: 1.7-34.6) among non-survivors. Among all hospitalized confirmed cases, censoring-adjusted probabilities of ICU admission and mortality were 41.9% (95% confidence interval: 34.1-51.4%) and 17.8% (14.3-22.2%), respectively, and higher among men than women. We estimated *R*_*E*_ was 1.43 (1.17-1.73), 2.09 (1.63-2.69), and 1.47 (0.07-2.59) in Northern California, Southern California, and Washington, respectively, for infections acquired March 1, 2020. *R*_*E*_ declined to 0.98 (0.76-1.27), 0.89 (0.74-1.06), and 0.92 (0.05-1.55) respectively, for infections acquired March 20, 2020.

**Conclusions:** We identify high probability of ICU admission, long durations of stay, and considerable mortality risk among hospitalized COVID-19 cases in the western United States. Reductions in *R*_*E*_ have occurred in conjunction with implementation of non-pharmaceutical interventions.

## INTRODUCTION

Months after its emergence in central China, the novel coronavirus SARS-CoV-2 has become pandemic, with cases of 2019 coronavirus disease (COVID-19) reported in nearly all countries.^1^ A total of 525,704 cases and 20,486 deaths were reported in the United States as of April 12, 2020, representing the greatest total of any country.^2^ Surges in COVID-19 cases have overwhelmed the capacity of hospital and healthcare systems in certain regions of the US, mirroring significant disruption witnessed in other countries.^3–5^ However, the epidemiology of COVID-19 in the United States remains poorly described, including clinical parameters of disease progression, the risk of intensive care unit (ICU) admission and death by patient age and sex, and the duration of hospital stay. As such, efforts to forecast trajectories of the epidemic to guide planning and response in the United States and other high-income settings have relied almost entirely on data from China to inform these parameters,^6^ which may not adequately reflect clinical circumstances elsewhere.

Metropolitan areas of the western United States were among the first to report COVID-19 importations and domestically-acquired cases. Washington state announced the first imported case of COVID-19 in the United States on January 21, 2020, while community transmission of COVID-19 has been identified since late February in Northern California. The vast majority of cases in California are concentrated in counties surrounding the San Francisco Bay in Northern California, and Los Angeles in Southern California. These regions were also among the first to implement intensive public health interventions aiming to curtail transmission. Social distancing recommendations for vulnerable populations were issued in San Francisco on March 6, 2020, and large gatherings were banned in Washington on March 11. Large-scale stay-at-home orders were implemented March 17 for the six counties of the San Francisco Bay area, and statewide in California and Washington on March 19 and March 24, respectively. Understanding the impact of these interventions is of crucial importance to inform their broader use, and may help to account for regional differences in the severity of COVID-19 epidemics across the country.^7^

To inform the epidemiology of COVID-19 in these regions, we analyzed healthcare data covering all hospitalized COVID-19 cases within the cohort of 9,596,321 individuals receiving comprehensive, integrated care from Kaiser Permanente (KP) healthcare systems in Northern California (KPNC), Southern California (KPSC), and Washington state (KPWA).

## METHODS

### Study design

The KPNC, KPSC, and KPWA systems deliver fully integrated healthcare to diverse membership cohorts generally resembling the commercially-insured populations of the surrounding geographic areas.^8–10^ We analyzed clinical and administrative data captured from all KP members who had been hospitalized within these KP care delivery systems with COVID-19 laboratory or clinical diagnoses at any recorded healthcare encounter by April 9, 2020. We considered patients to be clinically confirmed if no positive test result was available and diagnosis codes included any of the six diagnoses listed in **Table S1**. Clinical diagnoses were overruled by negative laboratory test results. Encounters included hospital, outpatient, and telehealth visits as well as uses of laboratory diagnostic services. We limited hospitalizations to acute inpatient admissions occurring between 14 days before to 28 days after the first encounter resulting in a COVID-19 diagnosis. Because only the most recent clinical encounter resulting in a COVID-19 diagnosis was available for KPWA patients, we considered COVID-19 hospitalizations to include those for which admission or discharge occurred within 30 days of the most recent COVID-19 encounter. Observational admissions were excluded. For the KPNC and KPSC cohorts, hospitalization events were sourced from electronic medical records and outside medical claims. Hospitalizations in the KPWA cohort were sourced from a centralized admissions database.

Patients discharged at the end of their most recent hospitalization were considered to be survivors if they did not die by April 9. As described below, we used competing-risk parametric survival methods to account for censoring of observations from currently-hospitalized patients for all analyses. Available data for patients included dates of COVID-19 clinical encounters, patient age, sex, dates of hospitalization, total duration of hospital stay, duration of ICU stay, ultimate clinical disposition (for completed hospitalizations only), and COVID-19 diagnostic tests performed in any setting as well as test results.

### Statistical analysis

#### Incidence estimation

We estimated daily and cumulative incidence of COVID-19 hospitalization within the full cohort population and within 10-year age strata for each of the KPNC, KPSC, and KPWA systems. Hospitalized case line lists included individuals with completed as well as ongoing hospitalizations as of April 9, 2020.

#### Duration of hospitalization

To inform planning, we aimed to infer distributions of the duration of hospitalization among all hospitalized patients and among survivors and non-survivors, and the distribution of the duration of ICU stay among patients receiving intensive care. For these analyses, we used the CFC package^11^ in R (version 1.1.463; R Foundation for Statistical Computing; Vienna, Austria) to fit age-adjusted Weibull competing risk models. Outcomes included discharge, mortality, or right censoring (for ongoing hospitalizations).

Duration of ICU stay was available only within the subset of patients with completed hospitalizations. For unbiased inference of the duration of ICU stay, we resampled observations from patients’ conditional distribution of ICU stay lengths, given their duration of hospitalization, age group, and survivor or non-survivor status. We defined sampling weights according to the (unconditional) distribution of total hospitalization durations across these patient strata, estimated as described above. We fitted Weibull distribution parameters to the resampled data via maximum likelihood to reconstruct unbiased distributions of total ICU stay.

#### Probability of ICU admission and mortality

We used a similar approach to correct for censoring of recent hospitalizations when estimating age- and sex-specific probabilities of ICU admission and death among hospitalized patients. Using generalized linear models with a Poisson link function, we estimated the conditional probability of each outcome among patients with completed hospitalizations given the duration of hospital stay, age, and sex, accounting for all two-way interactions among these covariates needed to minimize values of the Bayesian Information Criterion. We then integrated the estimated conditional probabilities of each outcome over all durations of hospitalization, weighted by the probability distribution of hospitalization durations within each age and sex stratum.

#### Transmission dynamics

We used hospitalization data to estimate cumulative numbers of infections over time, along with time-specific values of the effective reproductive number (*R*_*E*_), which describes the number of secondary infections resulting from infections acquired on a given day.^12^

We aimed to estimate cumulative infections by sampling the date of SARS-CoV-2 acquisition for each hospitalized case, and the number of infections occurring within the same age group on the same date as each case. We assumed time from infection to hospitalization was distributed according to the sum of random draws from fitted distributions of the time from infection to symptoms onset (incubation period) and the time from symptoms onset to hospitalization. We inferred the distribution of the incubation period by sampling from previous parameterizations of its length based on independent data sources from multiple countries^13–16^ (weighted by the number of subjects in each of these studies) and by fitting a Weibull distribution to the resulting pooled sample via maximum likelihood. In addition, we fitted a Gamma distribution of the time from symptoms onset to hospitalization in a previous study^17^ by minimizing summed squared errors relative to the reported mean and interquartile range. By drawing samples of infection times for each hospitalized case, we reconstructed the distribution of infection times for each patient hospitalized by April 9, 2020. In order to account for right-censoring of infections that were not hospitalized by this date, we divided the number of observed hospitalized infections estimated to have been acquired each date by the proportion of those infections that would be expected to have admission by April 9, according to the cumulative distribution function of time from infection to hospitalization.

To estimate the number of infections occurring for each hospitalization ascertained in our dataset, we used previously-reported age-specific estimates of the conditional probability of COVID-19 hospitalization, given SARS-CoV-2 infection. Aggregating estimates at ages 0-19y,^18^ we fitted Beta distributions minimizing summed squared errors relative to the means and 95% credible intervals of the reported distributions.

To estimate *R*_*E*_ for new infections acquired each day in each cohort, we applied the method of Wallinga and Teunis^12^ to our reconstructed estimates of daily new infections, using the distribution of the serial interval to assign the probability of a transmission link between any two infections on differing days. Similar to the analysis described for incubation periods, we sampled from serial interval estimates from previous studies^13,19–21^ according to the number of subjects for whom data were available, and fitted a Weibull distribution to the sampled data by maximum likelihood. To correct for right censoring of transmission pairs, we divided estimates of *R*_*E*_ for infections acquired each day *t* by the cumulative distribution function of the serial interval evaluated over the period from day *t* to the end of the study.

## Ethics approval

Retrospective reviews of de-identified administrative data for this study were considered exempt, non-human subjects research by the KPNC, KPSC, and KPWA institutional review boards.

## RESULTS

### Patients

In total, 1277 members were hospitalized with confirmed COVID-19 diagnoses as of April 9, 2020, with 539, 664, and 74 belonging to the KPNC, KPSC, and KPWA cohorts, respectively (**Table 1**). The median age of cases across all three cohorts was 60 years, with a range of 1-103 years and 50% of patients between 47-72 years of age. Four (0.3%) patients were under 20 years of age, 505 (39.5%) were ages 65 years or older, and 157 (12.2%) were ages 80 years or older; 725 (56.8%) were male. Laboratory confirmation of COVID-19 diagnosis was available for 1171 (91.7%) hospitalized patients as of April 9, 2020.

**Table 1:**
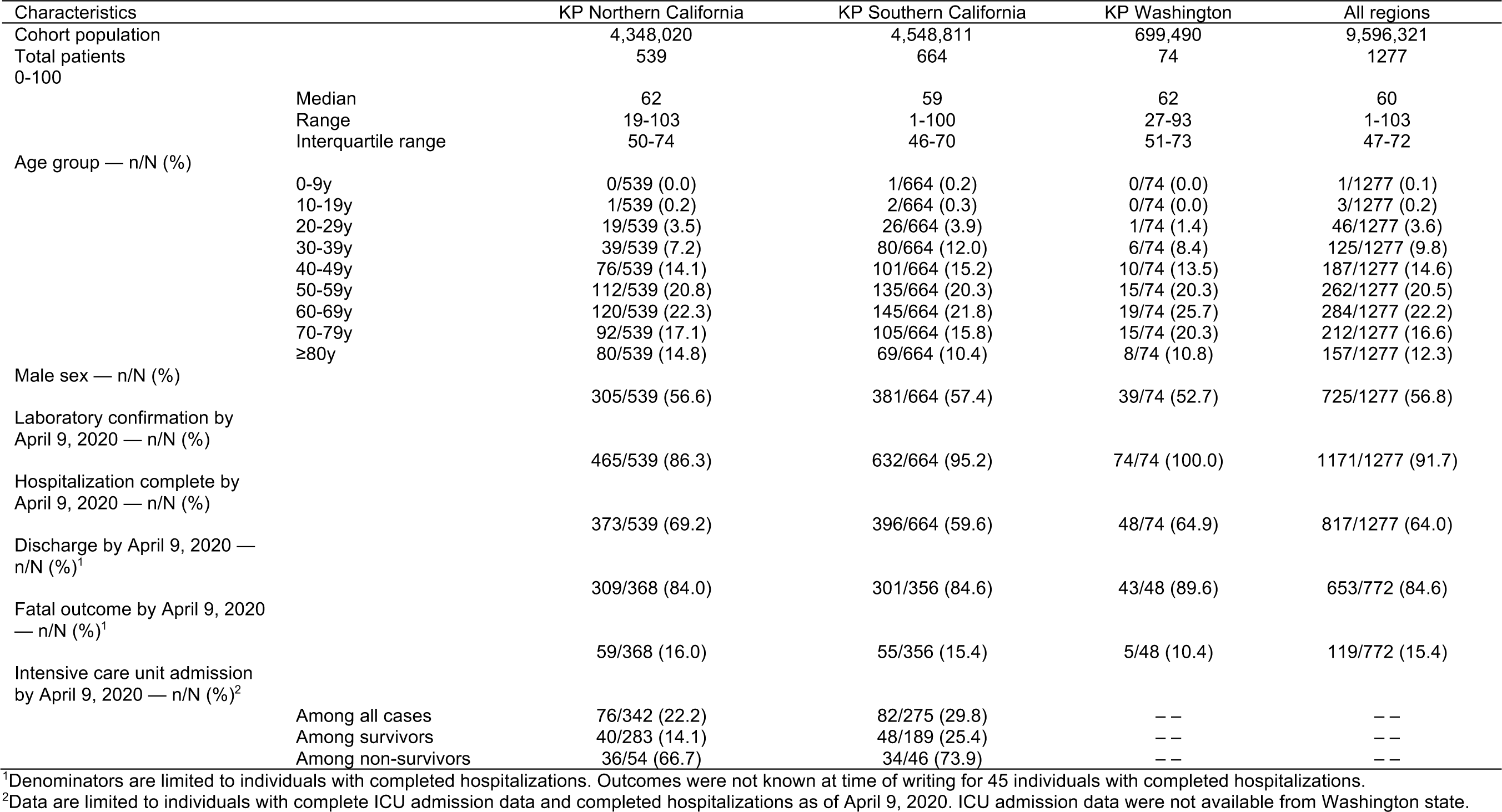
Characteristics of confirmed hospitalized COVID-19 cases.

Hospitalizations were complete for 817 (64.0%) individuals, among whom disposition data were complete for 772. Among all patents with completed hospitalizations and outcomes recorded, 119 (15.4%) were deceased by April 9, 2020. Data on ICU admission were available for 617 individuals (only those in the KPNC and KPSC cohorts), among whom 158 were admitted to ICU.

### Incidence of COVID-19 hospitalization

For the period ending April 9, 2020, we estimated the cumulative incidence of COVID-19 hospitalization within the KPNC, KPSC, and KPWA cohorts to be 12.4, 14.6, and 10.6 per 100,000 individuals (**Figure 1**). Incidence increased with age, reaching 61.0, 55.2, and 37.4 hospitalizations per 100,000 individuals ages ≥80 years in each of the three regions, respectively. Daily rates of hospitalization increased to the highest levels on March 26, 2020 in KPNC, and April 1 and 8 in KPSC and KPWA, respectively, with 6.4, 9.5, and 8.6 new daily admissions per million cohort members.

**Figure 1:**
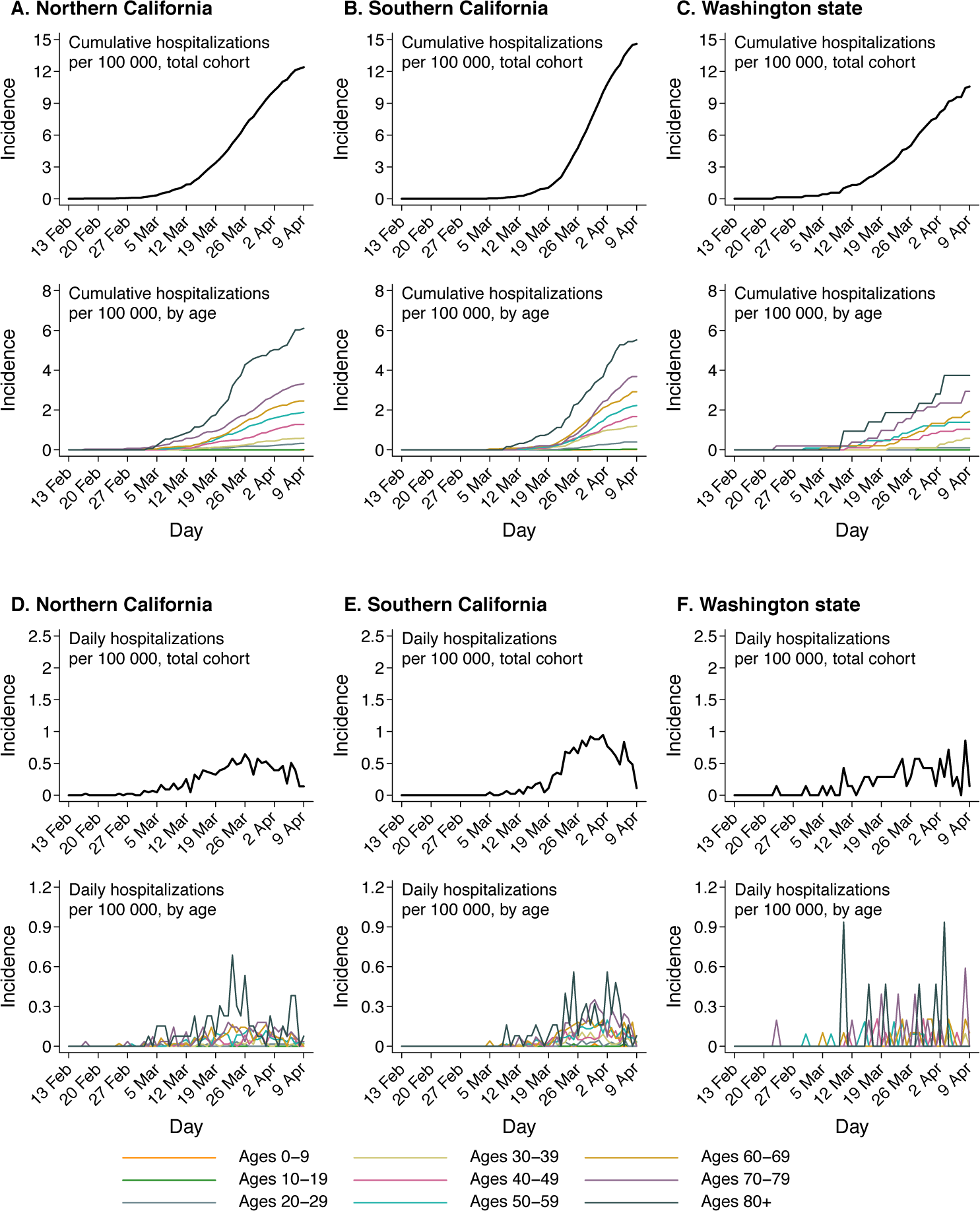
Cumulative and daily incidence of first COVID-19 hospitalization. We illustrate cumulative (**A-C**) and daily (**D-F**) incidence of first acute inpatient admission with confirmed COVID-19 diagnosis among all cohort members.

### Duration of hospital stay

Accounting for censoring of recent hospitalizations, we estimated the mean length of stay for all hospitalized patients to be 11.3 days, with 50% of hospitalizations lasting between 5.1-15.6 days and 95% lasting between 0.9-31.3 days (**Figure 2**; **Table S2**). Mean durations of stay for survivors and non-survivors were 10.7 and 13.7 days, with 95% of these patients staying 0.8-30.1 days and 1.7-34.6 days, respectively. The estimated mean duration of ICU stay was 8.1 days among patients receiving intensive care, with 50% of ICU stays lasting between 3.9-11.2 days and 95% lasting between 0.7-21.8 days.

**Figure 2:**
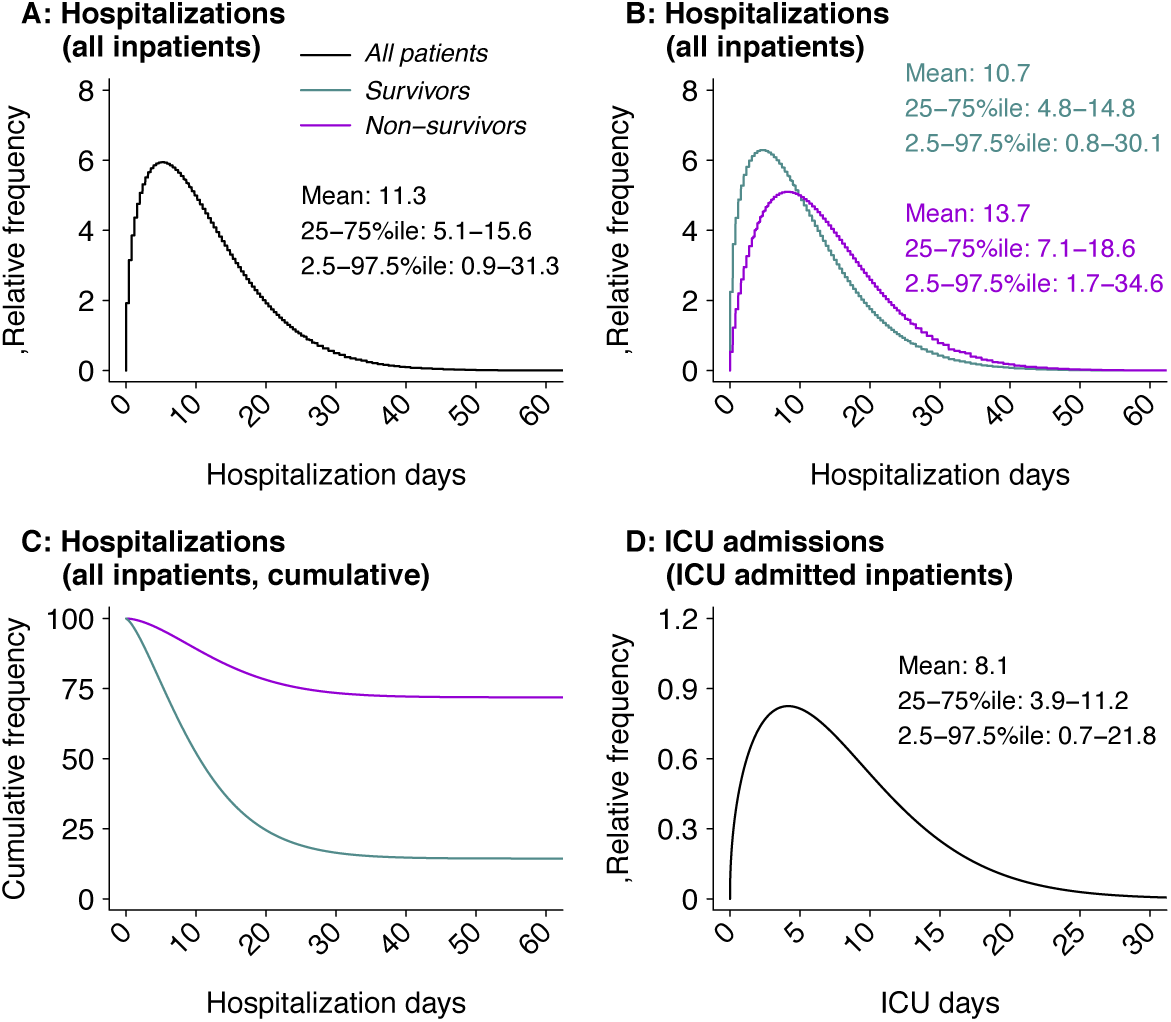
Durations of hospitalization among COVID-19 patients. In the top row, we illustrate the distributions of hospital length of stay for (**A**) all acute inpatient admissions; (**B**) acute inpatient admissions, stratified by clinical outcome; (**C**) time to discharge or death among all acute inpatient admissions; and (**D**) length of ICU stay for all inpatients admitted to ICU.

### Intensive care requirements and case fatality risk

Across all age groups and sexes (weighted by their representation among new admissions) and accounting for censoring of recent hospitalizations, we estimated 41.9% (34.1-51.4%) probability of ICU admission and 17.8% (14.3-22.2%) probability of death. Risk of death generally increased with age, while risk of ICU admission and death each tended to be higher among male than female patients (**Figure 3**). Risk of ICU admission appeared to increase with age among men only, with estimates ranging from 44.6% (26.4-76.0%) at ages 20-29 years to 70.7% (48.2-100.0%) at ages 70-79 years.

**Figure 3:**
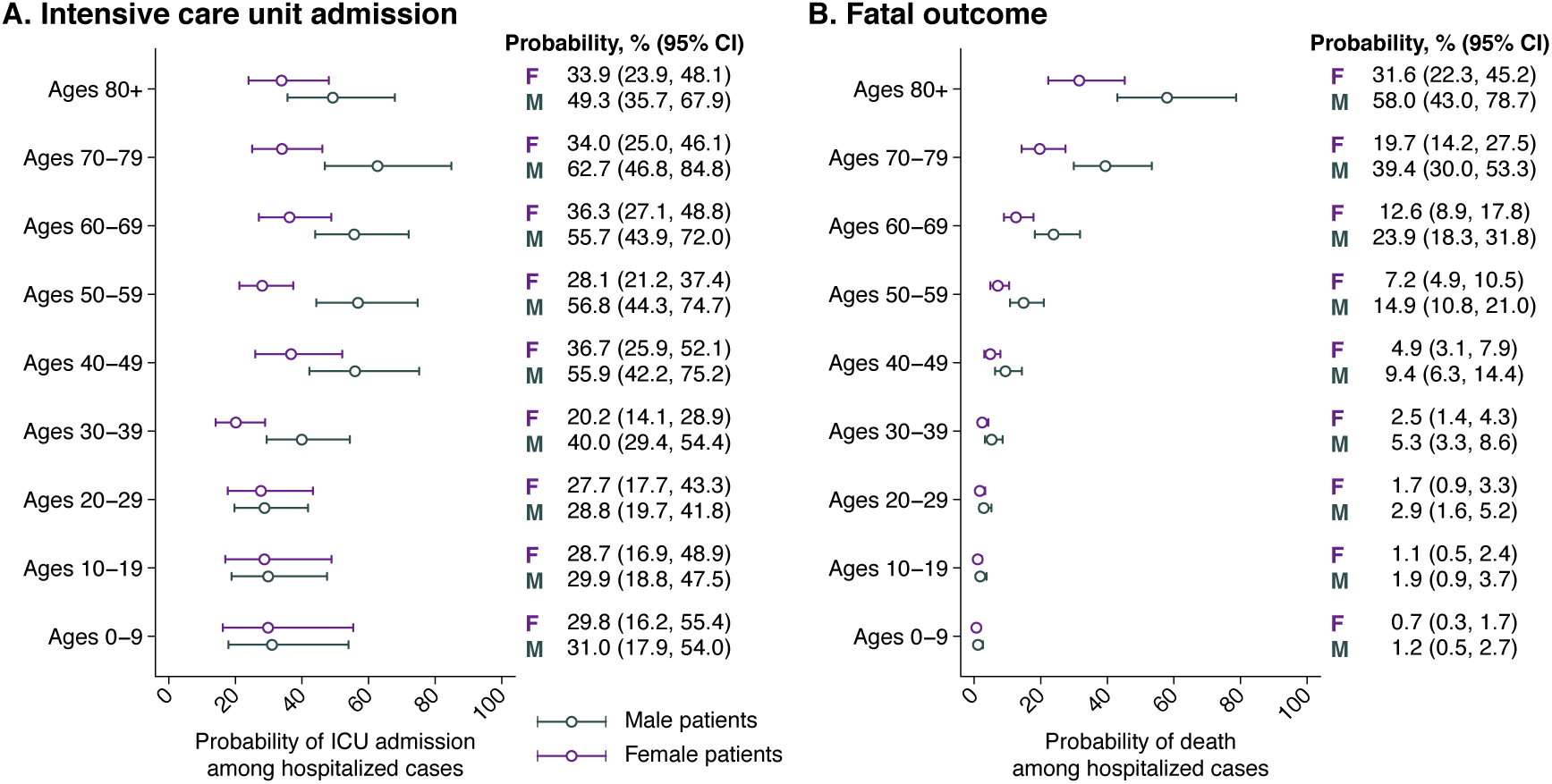
Probabilities of intensive care unit admission and mortality, by patient age and sex. We age- and sex-stratified probabilities of (**A**) intensive care unit admission and (**B**) mortality for male and female patients within 10-year age strata. Numerical estimates are indicated alongside plotted values. We obtain estimates using parametric (Weibull) survival models to account for censoring of observations among incomplete hospitalizations.

**Figure 4:**
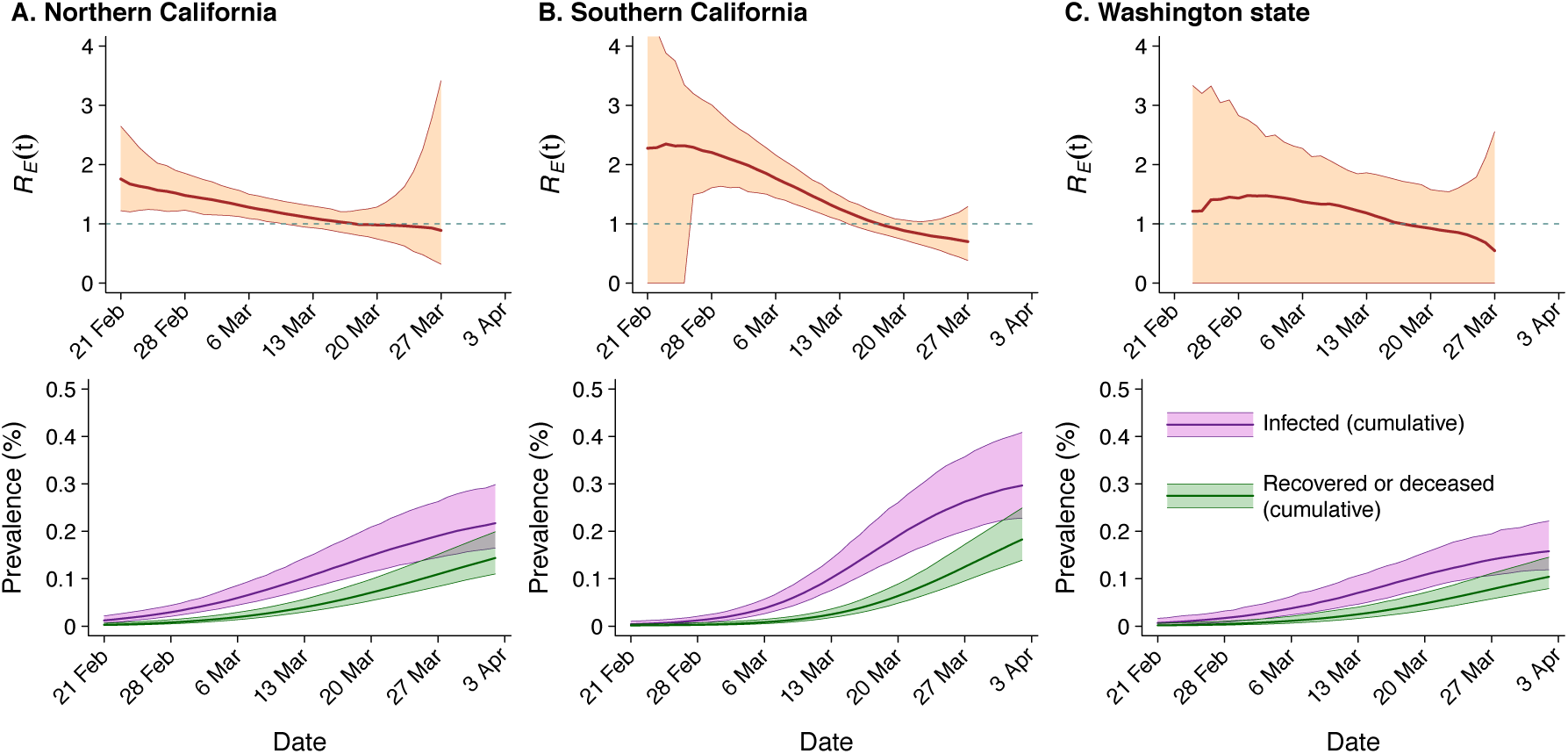
Dynamics of SARS-CoV-2 transmission in the cohort populations inferred from hospitalization data. We illustrate estimates of the effective reproductive number for infections acquired on day *t, R*_*E*_(*t*), describing the number of secondary infections each individual who acquired infection on day *t* would be expected to cause, for (**A**) Northern California, (**B**) Southern California, and (**C**) Washington state. Underneath, we plot estimates of the cumulative proportion of the population infected over time, and the proportion of the population that is deceased or recovered following previous infection. Shaded regions around point estimates (lines) indicate 95% confidence intervals.

### Transmission dynamics

Aggregating data across previous studies, we estimated a mean serial interval (the time between onset of an index case and onset of a secondary case) of 5.4 days (0.5-14.6), with a median length of 4.6 days. Our estimates of *R*_*E*_ indicated that individuals acquiring infection on March 1, 2020 were expected to cause an average of 1.43 (1.17-1.73), 2.09 (1.63-2.69), and 1.47 (0.07-2.59) secondary cases in Northern California, Southern California, and Washington state, respectively. Those acquiring infection on March 20, were expected to cause 0.98 (0.76-1.27), 0.89 (0.74-1.06), and 0.92 (0.05-1.55) secondary infections in the same settings.

We estimated a mean interval of 13.5 days (4.8-27.9) between infection and hospitalization for cases that would ultimately be hospitalized (**Table S3**). Accounting for the ratio of total infections to hospitalized cases, and for censoring of infections not yet hospitalized, we estimated the cumulative incidence of infection within the KPNC, KPSC, KPWA cohorts was 2.2 (1.7-3.1), 3.0 (2.3-4.1), and 1.6 (1.2-2.2) per 1000 individuals as of April 3, 2020.

## DISCUSSION

Our study provides an early assessment of the incidence and clinical profile of hospitalized COVID-19 cases among insured persons residing in the western United States and receiving comprehensive, integrated healthcare from KP health plans. Cumulative incidence of hospitalized COVID-19 cases grew exponentially within these managed care cohorts in Northern California, Southern California, and Washington state from late February through mid-March, 2020. The highest rates of incidence occurred among older adults, with 50% of hospitalizations occurring among adults ages ≥60 years and 25% of cases among adults ages ≥72 years. Consistent with differences in incidence of hospitalization, older patients experienced higher risks of ICU admission (among men) and death. Long durations of hospitalization, in particular among non-survivors, indicate the potential for substantial healthcare burden associated with management of severe COVID-19 cases.

In many respects, our findings are in agreement with observations in other settings, where older and male patients were more likely than younger or female patients to be admitted to the ICU and to die.^17,22–24^ The 11-day average duration of stay for hospitalized patients is consistent with observations in China.^22^ However, we estimated a 14-day average duration of stay among non-survivors, whereas non-survivors had a shorter length of hospitalization in China (7.5 days) than survivors.^22^ This difference may reflect, among other factors, alternative approaches to extending end-of-life care in the two settings. These findings have important ramifications for anticipating clinical needs. A widely-used model projecting clinical resource needs in the United Kingdom and United States assumes an 8 day mean duration of hospitalization for most patients.^6^ Similarly, our finding that 42% of hospitalized patients received intensive care is higher than the 30% estimate used for modeling based on observations in China.^6^

Notably, our hospitalized patient population was older on average than those hospitalized in China,^17,22,25,26^ where the median age was 50-57 years and only 25% were ages 68 and older. A small, single-center case series in Seattle, Washington reported an mean age of 70 years among ICU cases, consistent with the patient profile in our study.^27^ This observation of older ages in hospitalized US patients may relate to underlying population demographic structure as well as differences in health status and risk factor prevalence in US populations versus those of other settings.

Our estimates suggest *R*_*E*_ declined to a range near 1 amid the implementation of social distancing interventions, in line with declining increases in the incidence rate of new COVID-19 inpatient admissions. These reductions precede large-scale implementation of social distancing in the study regions. It should be noted that our daily *R*_*E*_(*t*) estimates describe transmission resulting from infections acquired each day *t*, rather than those transmitting on each day *t*. Because most individuals begin transmitting >4 days after acquiring infection, declines in *R*_*E*_ values are expected to precede dates of implementation of interventions that would affect transmission during individuals’ infectious periods. Individuals may have also taken precautionary measures to limit risk of acquiring or transmitting infection prior to implementation of stay-at-home orders. Similar observations have been reported in a study of transmission dynamics in King County, Washington.^28^ As our method propagates uncertainty in cumulative infection estimates based on time to hospitalization as well as the proportion of infected individuals experiencing symptoms, our approach does not aim to provide precise estimates of changes in *R*_*E*_ associated with interventions implemented on particular dates.

Our study has limitations. For this sample of 1277 hospitalized patients, we did not conduct a detailed review of medical records. As such, we do not address presenting characteristics of hospitalized patients and their association with demographic characteristics, length of hospital stay, or clinical outcome. Limited availability of laboratory testing in early phases of the US outbreak may have hindered ascertainment of sporadic cases in January and early February, 2020; because all persons under investigation for COVID-19 may not have received testing, our estimates of disease incidence should be interpreted as lower bounds. Near real-time hospitalization data may be missing for a modest subset of cases admitted to hospitals not owned by Kaiser Permanente, which would result in lagged reporting. In estimating transmission dynamics and cumulative infections, we relied on data from other settings to infer COVID-19 natural history parameters including the proportion of symptomatic infections requiring hospitalization, the serial interval, and the time from infection to hospitalization. Increases in the proportion of cases ascertained at later phases of the outbreak would be expected to increase *R*_*E*_ estimates over time, contrary to the trend we observed. Last, our estimation of *R*_*E*_ required an assumption that the KPNC, KPSC, and KPWA cohorts transmit among each other (or among epidemiologically similar individuals residing in the same areas). Within these regions, individuals receiving healthcare from KP health plans may be wealthier than those without commercial insurance. Economic security and employment type may impact individuals’ ability to comply with stay-at-home orders,^29^ meaning our estimates of transmission dynamics may not describe circumstances for other populations, including socioeconomically vulnerable groups. Despite this limitation, our use of data on hospitalized cases in a prospectively-followed cohort, receiving care within a unified healthcare delivery system, overcomes inconsistencies affecting *R*_*E*_ estimates from syndromic surveillance of milder COVID-19 cases across care providers and jurisdictions.^30^

The considerable length of stay among hospitalized cases in our study indicates that unmitigated transmission of SARS-CoV-2 poses a threat to US hospital capacity, consistent with observations in Italy and other high-resource settings^3^ as well as recent experience in New York.^2^ Our estimates of cumulative infections suggest the western United States remains far from reaching a herd immunity threshold. Although current social distancing measures have provided a crucial stopgap in reducing transmission and protecting healthcare systems to date,^29^ hospitals should ensure capacity to manage COVID-19 cases that will continue occurring in the coming months in a manner that is responsive to changes in social distancing or other pandemic-mitigating measures.

## Data Availability

Patient-level healthcare data are not publicly available. Code to reproduce technical aspects of the analysis is available from the corresponding author by request.

## ACKNOWLEDGMENTS

We thank the following individuals for their role in organization of data capture and preparation for analyses: Alvina Sundang, MBA, The Permanente Federation, Kaiser Permanente; Jay Robles, The Care Management Institute, Kaiser Permanente; and Lesley-Anne Myerscough, MSHS, Yumi Wong, Gina Gourd Bademian, KP Insight Washington, Kaiser Permanente. We thank the nationwide clinical, administrative, and operational teams of Kaiser Permanente for their effort in pandemic response.

The study was funded by Kaiser Permanente.

## DECLERATION OF INTERESTS

JAL, NPJ, and BLJ have received honoraria from Kaiser Permanente.

---

**Table S1:** Criteria used to ascertain laboratory- and clinically-confirmed COVID-19 cases.

| Outcome definition | Criteria |
| --- | --- |
| Laboratory-confirmed COVID-19 | Positive SARS-CoV-2 test result with or without diagnosis |
| Clinically-confirmed COVID-19 | Any of the of the following diagnoses without accompanying negative SARS-CoV-2 testing result: COVID-19 acute respiratory distress syndrome; COVID-19 pneumonia; COVID-19 acute bronchitis; COVID-19 lower respiratory infection; Asymptomatic COVID-19; Coronavirus disease 2019 (COVID-19) |

**Table S2:** Age-stratified estimates of hospital duration parameters.

| Age group | Duration of hospitalization, est. (2.5-97.5%ile); days |  |  |
| --- | --- | --- | --- |
|  | All hospitalizations | Survivors | Non-survivors |
| 0-9y | 6.8 (0.5-18.9) | 6.8 (0.5-18.9) | 8.0 (1.0-20.6) |
| 10-19y | 6.8 (0.5-18.9) | 6.8 (0.5-18.9) | 8.0 (1.0-20.6) |
| 20-29y | 7.6 (0.6-21.3) | 7.6 (0.6-21.3) | 9.0 (1.1-23.2) |
| 30-39y | 8.9 (0.7-24.7) | 8.8 (0.7-24.6) | 10.4 (1.2-26.8) |
| 40-49y | 9.8 (0.8-27.3) | 9.7 (0.8-27.1) | 11.5 (1.4-29.6) |
| 50-59y | 11.0 (0.9-30.3) | 10.8 (0.9-30.0) | 12.8 (1.5-32.7) |
| 60-69y | 12.2 (1.0-33.1) | 11.9 (1.0-32.6) | 13.9 (1.7-35.4) |
| 70-79y | 12.9 (1.2-34.2) | 12.3 (1.0-33.3) | 14.4 (1.8-36.0) |
| 80-89y | 12.8 (1.2-33.5) | 12.0 (1.0-32.3) | 14.0 (1.8-34.8) |
| All ages | 11.3 (0.9-31.3) | 10.7 (0.8-30.2) | 13.7 (1.7-34.6) |

Estimates are obtained by a fitting a multi-cause, competing risks model under the assumption of Weibull-distribute event times.

**Table S3:** Parameters obtained from other studies.

| Parameters |  | Distribution | Source |
| --- | --- | --- | --- |
| Time from infection to shedding | | Weibull($k = 5.983, \lambda = 1.455$) | refs.<sup>13,19-21</sup> |
| Time from shedding to symptoms | | Weibull($k = 0.294, \lambda = 0.14$) | refs.<sup>13-16</sup> |
| Time from symptoms onset to hospitalization | | Gamma($k = 5.078, \theta = 0.765$) | ref.<sup>17</sup> |
| Time shedding onset to clearance | | Exp($1/9$) | ref.<sup>6</sup> |
| Probability of hospitalization, given infection |  |  |  |
| | 0-9y | Beta($\alpha = 8.65, \beta = 1880.26$) | ref.<sup>18</sup> |
| | 10-19y | Beta($\alpha = 8.65, \beta = 1880.26$) | |
| | 20-29y | Beta($\alpha = 9.23, \beta = 767.30$) | |
| | 30-39y | Beta($\alpha = 8.43, \beta = 208.96$) | |
| | 40-49y | Beta($\alpha = 8.07, \beta = 159.93$) | |
| | 50-59y | Beta($\alpha = 7.85, \beta = 76.71$) | |
| | 60-69y | Beta($\alpha = 7.45, \beta = 48.25$) | |
| | 70-79y | Beta($\alpha = 7.01, \beta = 30.02$) | |
| | 80-89y | Beta($\alpha = 8.35, \beta = 32.67$) | |

## REFERENCES

1. World Health Organization. Coronavirus disease 2019 (COVID-19) Situation Report – 83. 2020 https://pers.droneemprit.id/covid19/. Accessed 12 April, 2020.

2. Centers for Disease Control and Prevention. Coronavirus disease 2019 (COVID-19) cases in US. https://www.cdc.gov/coronavirus/2019-ncov/cases-updates/cases-in-us.html. Accessed 12 April, 2020.

3. Remuzzi A, Remuzzi G. COVID-19 and Italy: what next? Lancet 2020; 2: 10–3.

4. Tanne JH, Hayasaki E, Zastrow M, Pulla P, Smith P, Rada AG. Covid-19: how doctors and healthcare systems are tackling coronavirus worldwide. BMJ 2020. DOI:10.1136/bmj.m1090.

5. Khan S, Nabi G, Han G, et al. Novel coronavirus: how things are in Wuhan. Clin Microbiol Infect 2020. DOI:10.1016/j.cmi.2020.02.005.

6. Ferguson NM, Laydon D, Nedjati-Gilani G, et al. Impact of non-pharmaceutical interventions (NPIs) to reduce COVID-19 mortality and healthcare demand. Imperial College, London 2020. DOI:10.25561/77482.

7. Chowell G, Mizumoto K. The COVID-19 pandemic in the USA: what might we expect? Lancet 2020. DOI:10.1016/S0140-6736(20)30743-1.

8. Koebnick C, Langer-Gould AM, Gould MK, et al. Sociodemographic characteristics of members of a large, integrated health care system: comparison with US Census Bureau data. Perm J 2012;16:37–41.

9. Gordon NP. Similarity of the adult Kaiser Permanente Membership in Northern California to the insured and general population in Northern California: Statistics from the 2009 California Health Interview Survey. Intern Div Res Rep 2012; 1–15.

10. Saunders KW, Davis RL, Stergachis A. Group health cooperative. Pharmacoepidemiology 2006; 223–39.

11. Mahani AS, Sharabiani MTA. Bayesian, and non-bayesian, cause-specific competing-risk analysis for parametric and nonparametric survival functions: The R Package CFC. J Stat Softw 2019. DOI:10.18637/jss.v089.i09.

12. Wallinga J, Teunis P. Different epidemic curves for severe acute respiratory syndrome reveal similar impacts of control measures. Am J Epidemiol 2004. DOI:10.1093/aje/kwh255.

13. Tindale L, Coombe M, Stockdale JE, et al. Transmission interval estimates suggest pre-symptomatic spread of COVID-19. medRxiv 2020. DOI:10.1101/2020.03.03.20029983.

14. Backer JA, Klinkenberg D, Wallinga J. Incubation period of 2019 novel coronavirus (2019-nCoV) infections among travellers from Wuhan, China, 20-28 January 2020. Euro Surveill 2020. DOI:10.2807/1560-7917.ES.2020.25.5.2000062.

15. Lauer SA, Grantz KH, Bi Q, et al. The incubation period of coronavirus disease 2019 (COVID-19) from publicly reported confirmed cases: Estimation and application. Ann Intern Med 2020. DOI:10.7326/M20-0504.

16. Linton NM, Kobayashi T, Yang Y, et al. Incubation period and other epidemiological characteristics of 2019 novel coronavirus infections with right truncation: a Statistical analysis of publicly available case data. J Clin Med 2020. DOI:10.3390/jcm9020538.

17. Wang D, Hu B, Hu C, et al. Clinical characteristics of 138 hospitalized patients with 2019 novel coronavirus-infected pneumonia in Wuhan, China. JAMA 2020. DOI:10.1001/jama.2020.1585.

18. Verity R, Okell LC, Dorigatti I, et al. Estimates of the severity of coronavirus disease 2019?: a model-based analysis. Lancet Infect Dis 2020; 3099: 1–9.

19. Du Z, Xu X, Wu Y, Wang L, Cowling BJ, Meyers LA. Serial Interval of COVID-19 among Publicly Reported Confirmed Cases. Emerg Infect Dis J 2020; 26. DOI:10.3201/eid2606.200357.

20. Nishiura H, Linton NM, Akhmetzhanov AR. Serial interval of novel coronavirus (COVID-19) infections. Int J Infect Dis 2020. DOI:10.1016/j.ijid.2020.02.060.

21. Li Q, Guan X, Wu P, et al. Early transmission dynamics in Wuhan, China, of novel coronaviru-infected pneumonia. N Engl J Med 2020. DOI:10.1056/nejmoa2001316.

22. Zhou F, Yu T, Du R, et al. Clinical course and risk factors for mortality of adult inpatients with COVID-19 in Wuhan, China: a retrospective cohort study. Lancet 2020. DOI:10.1016/S0140-6736(20)30566-3.

23. Grasselli G, Zangrillo A, Zanella A, et al. Baseline Characteristics and Outcomes of 1591 Patients Infected With SARS-CoV-2 Admitted to ICUs of the Lombardy Region, Italy. JAMA 2020; 2–9.

24. Onder G, Rezza G, Brusaferro S. Case-Fatality Rate and Characteristics of Patients Dying in Relation to COVID-19 in Italy. JAMA 2020; 2019: 2019–20.

25. Qian G-Q, Yang N-B, Ding F, et al. Epidemiologic and Clinical Characteristics of 91 Hospitalized Patients with COVID-19 in Zhejiang, China: A retrospective, multi-centre case series. QJM 2020. DOI:10.1093/qjmed/hcaa089.

26. Liu K, Fang Y-Y, Deng Y, et al. Clinical characteristics of novel coronavirus cases in tertiary hospitals in Hubei Province. Chin Med J 2020. DOI:10.1097/cm9.0000000000000744.

27. Arentz M, Yim E, Klaff L, et al. Characteristics and outcomes of 21 critically ill patients with COVID-19 in Washington state. JAMA 2020. DOI:10.1001/jama.2020.4326.

28. Thakkar N, Burstein R, Hu H, Selvaraj P, Klein D. Social distancing and mobility reductions have reduced COVID-19 transmission in King County, WA. Institute for Disease Modeling; 2020. https://covid.idmod.org/data/Social_distancing_mobility_reductions_reduced_COVID_Seattle.pdf. Accessed 12 April, 2020.

29. Lewnard JA, Lo NC. Scientific and ethical basis for social-distancing interventions against COVID-19. Lancet Infect Dis 2020. DOI:10.1016/S1473-3099(20)30190-0.

30. Lipsitch M, Swerdlow DL, Finelli L. Defining the Epidemiology of Covid-19 - Studies Needed. N Engl J Med 2020. DOI:10.1056/NEJMp2002125.

